# Limited specificity of SARS-CoV-2 antigen-detecting rapid diagnostic tests at low temperatures

**DOI:** 10.1101/2021.02.01.21250904

**Authors:** Verena Haage, Andres Moreira-Soto, Jilian A. Sacks, Victor Corman, Christian Drosten, Felix Drexler

## Abstract

SARS-CoV-2 antigen-detecting rapid diagnostic tests (Ag-RDTs) are available within and outside of health care settings to enable increased access to COVID-19 diagnosis. These environments include provisional testing facilities, lacking temperature control; as outside temperatures fall, recommended testing temperatures cannot be guaranteed. We report impaired specificity in two out of six Ag-RDTs when used at 2-4°C, indicating that testing in cold settings might cause false-positive results potentially entailing unwarranted quarantine assignments and incorrect incidence estimates.

## Introduction

SARS-CoV-2 antigen-detecting tests were introduced into the global market during late 2020 and are now widely used, in both the global north and south. Many regions located in the northern hemisphere such as the United States or Europe are currently severely affected by the second wave of the COVID-19 pandemic (1, 2). To manage testing demand many physicians, healthcare and public health systems have opened external testing stations such as ‘diagnostic streets’ or drive-through facilities in urban settings (3). In Berlin, a German metropolis with a population of about 3.5 million inhabitants, several outside testing stations in tents or pavilions are in use, produced by manufacturers who offer special corona test cabins (4). These facilities are often of provisional nature, for example in the form of unheated tents. In the winter months, temperatures in Europe or the U.S. can range from −10°C to 10°C (5, 6), well below the recommended operating temperatures of most Ag-RDTs. Most manufacturers of SARS-CoV-2 Ag-RDTs specify storage conditions between 2-30°C, but stipulate that tests be equilibrated to room temperature (15-30°C) at the time of use to guarantee their performance. With temperatures around freezing point during the winter months, unheated testing facilities cannot always comply with these conditions. Moreover, as the national COVID-19 reference laboratory in Germany, we were contacted by physicians from two German metropolis both reporting an unusual high number of positive SARS-CoV-2 Ag-RDTs from outside testing facilities. To test whether operation of SARS-CoV-2 Ag-RDTs at low outside temperatures impacts their performance, we selected six (**Table 1**) out of the 139 currently commercially available SARS-CoV-2 Ag-RDTs (7) based on clinical performance data (8) and market availability, for validation of specificity when stored and/or operated at cold outside temperatures of 2-4°C.

**Table 1.** Overview of SARS-CoV-2 rapid antigen tests included in the study

| ID | Test | Manufacturer | Lot No. |
| --- | --- | --- | --- |
| I | Panbio™ COVID-19 Ag Rapid Test | Abbott Laboratories | 41ADF012A |
| II | ActivXpress + COVID-19 Antigen Complete Testing Kit | Edinburgh Genetics | AG20200905 |
| III | COVID-19 Ag | Genedia | 643X2005 |
| IV | ichroma - COVID-19 Ag | Boditech Med | SRQHA27 |
| V | COVID-19 Antigen Rapid Test Kit | JOYSBIO (Tianjin) Biotechnology Co., Ltd. | 2020092409 |
| VI | SARS-CoV-2 Rapid Antigen Test | Roche Diagnostics* | QCO3020083<br>QCO390003I/Sub:I-2<br>QCO390011A/Sub:A-2 |

\* equals STANDARD Q COVID-19 Test by SD Biosensor, Inc.

## Materials and Methods

Tested conditions included: (a) storage and testing of Ag-RDTs at recommended conditions (15-30°C); (b) storage of tests at recommended conditions, pre-incubation of tests for 30mins to cold-temperatures (2-4°C) and testing at cold temperatures (2-4°C); and (c) and storage of tests at 2-4°C followed by testing at 2-4°C (**Figure 1A**). Analytical specificity was examined by testing for cross-reactivity with common cell culture-derived respiratory viruses including the ubiquitous human coronaviruses (HCoV) HCoV-229E (2.9×10^7^ copies/ml), HCoV-OC43 (1.0×10^6^ copies/ml), influenza virus A H1N1 (7.8×10^6^ copies/ml) and rhinovirus A (2.2×10^6^ copies/ml). Concentrations were selected according to the guidelines on analytical specificity testing for SARS-CoV-2 Ag-RDTs published by the German Federal institute for vaccines and biomedicines (9). 20µl of viral cell culture supernatant were added to proprietary lysis buffer or as an internal control, and 20µl of lysis buffer were directly applied to test cassettes for validation experiments. Tests were performed in duplicates, according to the test-specific supplier instructions for use and results were scored independently by two persons. We additionally tested ten healthy laboratory members who previously volunteered for a SARS-CoV-2 Ag-RDT validation study (8). Healthy volunteers were without symptoms of respiratory tract infection and tested negative for SARS-CoV-2 by RT-qPCR (10). All subjects received instructions on self-sampling, swabs were immediately dissolved in 1ml PBS and 20µl of PBS were added to proprietary buffer for testing. In a recent study, self-sampling was shown to be a reliable alternative to professional nasopharyngeal swabs for Ag-RDTs (11).

**Figure 1.**
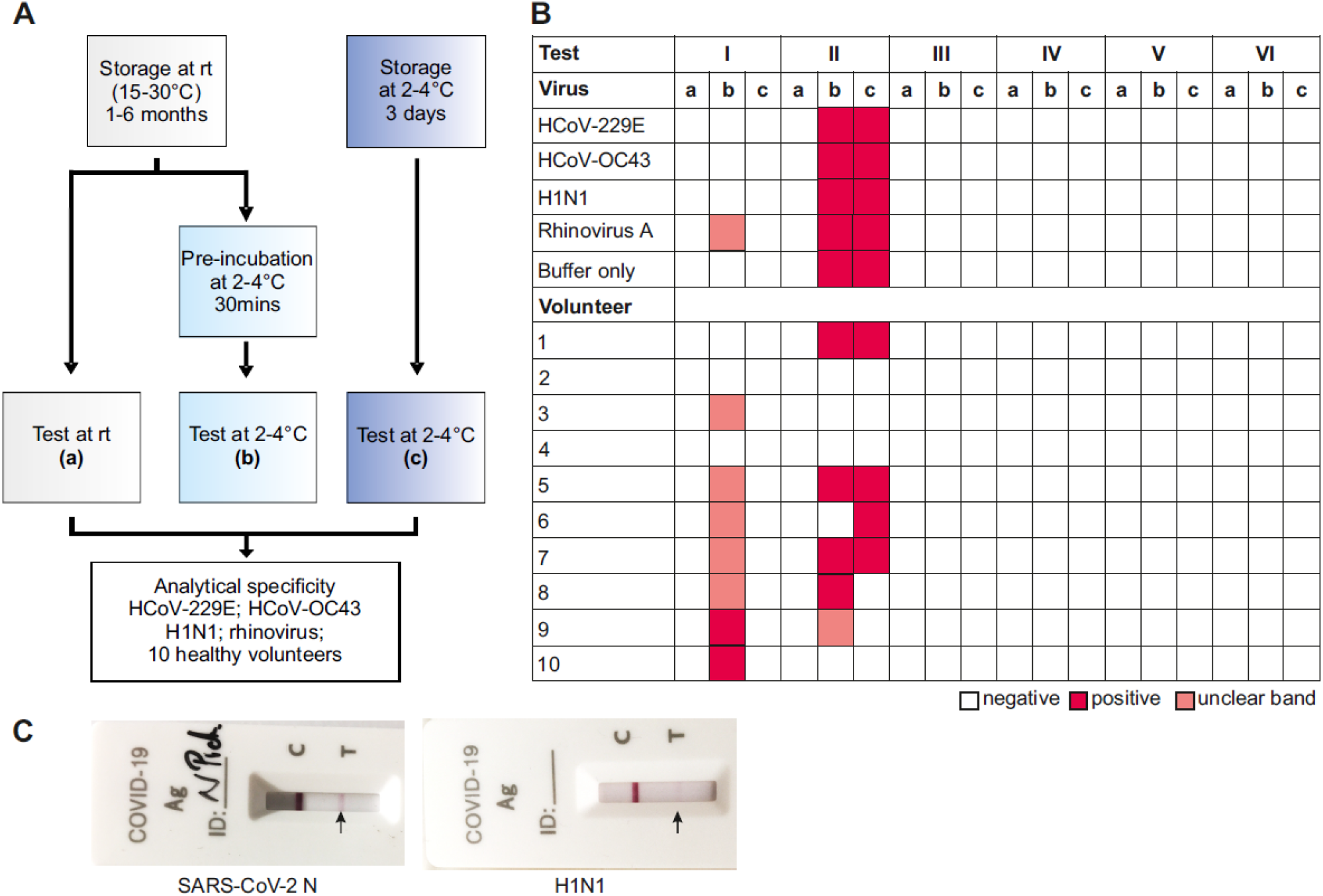
Experimental setup and results for specificity testing of SARS-CoV-2 Ag-RDTs. **A**. (a): storage and operation at recommended conditions; (b): recommended storage, pre-incubation at 2-4°C for 30mins and operation at 2-4°C; (c): storage and operation at 2-4°C. **B**. Specificity of SARS-CoV-2 Ag-RDTs decreases at low temperatures. (a): rt storage-rt testing; (b): rt storage – 2-4°C 30 min pre-incubation – 2-4°C testing; (c): 2-4°C storage – 2-4°C testing. red: positive; white: negative; salmon: weak band, result unclear. rt: room temperature. I: Abbott; II: ActivXpress; III: Genedia; IV: ichroma; V: JOYSBIO; VI: Roche. C. Example for observed cross-reactivity of the ActivXpress test with Influenza virus A H1N1 and SARS-CoV-2 nucleoprotein as positive control (SARS-CoV-2-N; 5µg/ml) when tested under condition (b): recommended storage, pre-incubation at 2-4°C for 30mins and operation at 2-4°C.

## Results

Two of the six SARS-CoV-2 Ag-RDTs showed impaired specificity (**Figure 1B**) when stored at room temperature, but exposed to 2-4°C for 30 minutes prior to testing at 2-4°C as cross-reactivity with common respiratory viruses, and false-positive results occurred in healthy volunteers in the form of weak, but clearly visible bands (**Figure 1C**). In one test (test I), unspecific reactivity was only observed upon short-term incubation at 2-4°C followed by test operation at 2-4°C, but not after long-term storage at 2-4°C. In contrast, the other test yielding unspecific results (test II) yielded almost identically unspecific results after both, short- and long-term storage at 2-4°C and operation at 2-4°C (**Figure 1B**). On the one hand, those data highlight differences between test devices. On the other hand, our results may hint at effects of relatively rapid temperature changes on some tests for unknown reasons, potentially including environmental factors such as condensation. Results were reproducible and functionality of tests was confirmed by determining their level of detection using serial dilutions of SARS-CoV-2 nucleoprotein (SARS-CoV-2-N) at recommended conditions as previously described (8).

## Discussion

Our study highlights that specificity of SARS-CoV-2 Ag-RDTs may be impaired when operating tests at temperatures that differ from recommended conditions (15-30°C), leading to false-positive results. These results were observed for only certain test brands including one of the Ag-RDT currently listed for emergency use by the World Health Organization (12, 13), highlighting that each test may need to be considered specifically and broader validation of temperature robustness of SARS-CoV-2 RDTs should be performed. Moreover, impaired sensitivity of SARS-CoV-2 Ag-RDTs at elevated temperatures has been reported previously (14), highlighting the importance of test operation at recommended temperatures. Of note, all tests studied here were shown to be highly specific when operated at recommended conditions in prior studies (8, 14), underlining that impaired specificity is not a test-intrinsic problem but owed to test operation under conditions beyond those defined by the manufacturer.

Our data imply that caution must be taken when offering SARS-CoV-2 Ag-RDT diagnostics in settings lacking temperature control, including diagnostic streets or drive-through testing stations in car parks (3). Especially under provisional conditions when minimally-trained or unexperienced staff perform diagnostic tests, the establishment of laboratory workflows and compliance with the conditions recommended by the manufacturer are vital to ensure accurate testing (15). Until January 14^th^ 2021, severity of regional lockdown measures in Germany is increased upon trespassing 7-day incidences of 200 per 100.000 inhabitants (16), highlighting the potential consequences of false-positive test results beyond potentially unwarranted individual quarantine assignments, if those results were reported to public health authorities without confirmation by a gold standard test such as RT-PCR (10).

## Data Availability

The authors confirm that the data supporting the findings of this study are available within the article.

## Funding

The study was partially supported by the Foundation for Innovative New Diagnostics (FIND), including procurement of some test kits. The findings and conclusions contained within are those of the authors and do not necessarily reflect positions or policies of FIND.

## Conflict of interest

The authors declare no conflict of interest.

## Notes

### Competing Interest Statement

The authors have declared no competing interest.

### Author Declarations

The testing of employees is part of an ongoing study on SARS-CoV-2 infection in employees under Charite ethical review board file number EA1/068/20.

